# Attributes and predictors of Long-COVID: analysis of COVID cases and their symptoms collected by the Covid Symptoms Study App

**DOI:** 10.1101/2020.10.19.20214494

**Authors:** Carole H. Sudre, Benjamin Murray, Thomas Varsavsky, Mark S. Graham, Rose S. Penfold, Ruth C. Bowyer, Joan Capdevila Pujol, Kerstin Klaser, Michela Antonelli, Liane S. Canas, Erika Molteni, Marc Modat, M. Jorge Cardoso, Anna May, Sajaysurya Ganesh, Richard Davies, Long H Nguyen, David A. Drew, Christina M. Astley, Amit D. Joshi, Jordi Merino, Neli Tsereteli, Tove Fall, Maria F. Gomez, Emma L. Duncan, Cristina Menni, Frances M.K. Williams, Paul W. Franks, Andrew T. Chan, Jonathan Wolf, Sebastien Ourselin, Tim Spector, Claire J. Steves

## Abstract

Reports of “Long-COVID”, are rising but little is known about prevalence, risk factors, or whether it is possible to predict a protracted course early in the disease. We analysed data from 4182 incident cases of COVID-19 who logged their symptoms prospectively in the COVID Symptom Study app. 558 (13.3%) had symptoms lasting >=28 days, 189 (4.5%) for >=8 weeks and 95 (2.3%) for >=12 weeks. Long-COVID was characterised by symptoms of fatigue, headache, dyspnoea and anosmia and was more likely with increasing age, BMI and female sex. Experiencing more than five symptoms during the first week of illness was associated with Long-COVID, OR=3.53 [2.76;4.50]. A simple model to distinguish between short and long-COVID at 7 days, which gained a ROC-AUC of 76%, was replicated in an independent sample of 2472 antibody positive individuals. This model could be used to identify individuals for clinical trials to reduce long-term symptoms and target education and rehabilitation services.

---

COVID-19 can manifest a wide severity spectrum from asymptomatic to fatal forms^1^. A further source of heterogeneity is the duration of symptoms, which could have considerable impact due to the huge scale of the pandemic. Hospitalised patients are well recognised to have lasting dyspnoea and fatigue in particular^2^, yet such patients constitute the ‘tip of the iceberg’ of symptomatic SARS CoV2 disease^3^. Few studies capture symptoms prospectively in the general population to ascertain with accuracy the duration of illness and the prevalence of long-lasting symptoms.

Here we report a prospective observational cohort study of COVID-19 symptoms in a subset of 4182 users of the COVID Symptom Study app meeting inclusion criteria (see online methods)^4,5^, compared to matched symptomatic test-negative controls. Briefly, the cases comprised individuals who reported testing positive for SARS-CoV2 by swab testing who started on the app “feeling physically normal” to be able to determine symptom onset. We compare cases with symptoms persisting over 28 days, LC28) and short duration (symptoms lasting less than 10 days, short-COVID). Our previous findings that clusters of symptoms predicted the need for acute care^6^ led us to hypothesize that persistent symptomatology in COVID-19 (Long-COVID) is associated with early symptom patterns which could be used to predict who might be affected. Figure 1 shows the duration of symptoms reported in COVID+ cases (orange) over-laid on age, sex and BMI matched negative-testing symptomatic controls (blue). The overall median symptom duration was 11 days (IQR[6;19]).

**Figure 1.**
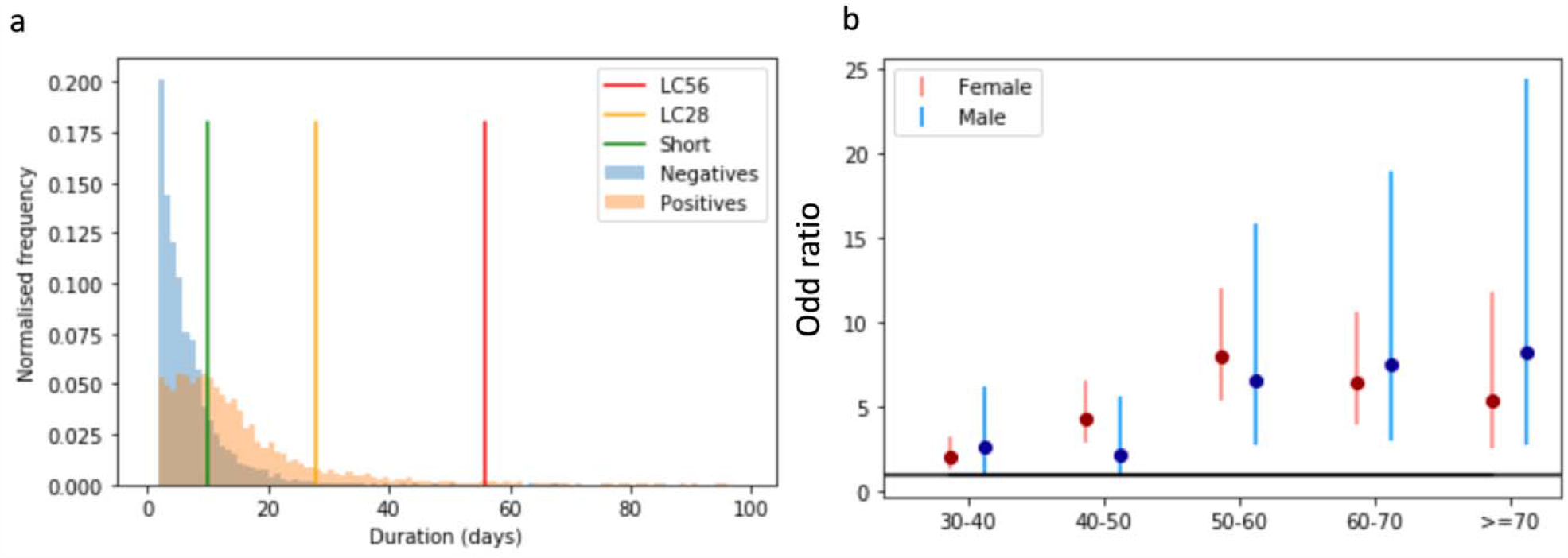
a) Distribution of duration of symptoms in COVID-19 – The coloured bars indicate the limits to define short, LC28 and LC56. The y-axis reports the normalised frequency of duration of symptoms. b) OR and 95% CI of LC28 with each successive decile compared to 20-30-year-olds

Of the 4182 COVID-19 swab positive users, 558 (13.3%) met the LC28 definition (Median 41, IQR[33,63]) of whom 189 (4.5%) met LC56, and 108 (2.6%) LC84. In contrast 1591 (38.0%) had short-COVID (median 6, IQR[4-8]). The proportions were comparable in three countries (LC28: GB 13.3%, USA 16.1%, Sweden 12.1% p=0.35; LC56: GB 4.7%, USA 5.5%, Sweden 2.5% p=0.07).

Table 1 summarises the descriptive characteristics of the study population stratifying by symptom/disease duration. Age was significantly associated with LC28, rising from 9.9% in 18-49 year-olds to 21.9% in those aged >=70 (p < 0.0005), with escalating OR by age decile (Figure 1b, Supplementary Table 2). LC28 disproportionately affected women (14.9%) compared to men (9.5%), although not in the older age-group. Long-COVID affected all socio-economic groups (assessed using Index of Multiple Deprivation), (Supplementary Figure 2). Individuals with Long-COVID were more likely to have required hospital assessment. Asthma was the only/unique pre-existing condition providing significant association with LC28 (OR=2.14 [1.55-2.96]).

**Table 1.** Characteristics of Individuals with COVID-19 by duration of symptoms, compared to matched sample testing negative for COVID-19. In statistical comparison, the short COVID group is the reference.

|  | Positive PCR test |  |  |  | Matched Negative sample |
| --- | --- | --- | --- | --- | --- |
|  | Short (<10 days) | LC28 (>=28 days)<br>(including LC56) | LC56 (>=56 days) | Overall |  |
| Number | 1591 | 558 | 189 | 4182 | 4182 |
| GB/SE/US<br>[numbers, %] | 1365 / 139 / 87<br>85.8 / 8.7 / 5.5 | 466/57/35<br>83.5 / 10.2 / 6.3 | 165/12/12<br>87.3 / 6.3 / 6.3 | 3491 / 473 / 218<br>83.5 / 11.3 / 5.2 | 3882/131/169<br>92.8/3.1/4.1 |
| Male (%) | 32.7 | 20.3*** | 16.9* | 28.5 | 28.5 |
| Age (years)<br>[median, IQR] | 38 [29;49] | 50 [39;57]*** | 52 [43;59]*** | 42 [32;53] | 42 [32;53] |
| Age group<br>(18-49/ 50-69/<br>>70) | 1122/331/38<br>75.3 / 22.2 / 2.5 | 259/262/24<br>47.5 / 48.1 / 4.4 | 69/96/11<br>39.2 / 54.5 / 6.3 | 2627/1195/96<br>62.8 / 28.6 / 2.3 | 2821 / 1264/97<br>67.5 / 30.2 / 2.3 |
| Obese (%) | 23.8 | 27.6* | 26.5 | 26.3 | 26.4 |
| BMI (kg/m<sup>2</sup>)<br>[median, IQR] | 25.5 [22.7;29.7] | 26.1 [23.3;30.5] | 25.9[23.3;30.5] | 25.9[23.3;30.3] | 25.9 [23.0;30.3] |
| Asthma (%) | 7.7 | 15.8*** | 18.0*** | 10.0 | 13.7 |
| Lung disease<br>(%) | 12.8 | 16.5** | 15.9 | 13.6 | 13.7 |
| Diabetes (%) | 3.0 | 3.9 | 5.8* | 2.9 | 2.8 |
| Heart (%) | 1.7 | 3.2** | 4.8** | 1.9 | 1.7 |
| Kidney (%) | 0.5 | 0.9 | 0.5 | 0.6 | 0.6 |
| IMD (median decile - IQR) | 7 [4;9] | 7 [5;9] | 7 [5;9] | 7 [4;9] | 7[5;9]*** |
| IMD quintiles | 64/75/334/132/634<br>5.2/6.1/27.0/10.7/51.2 | 23/23/86/49/240<br>5.5/5.5/20.4/11.6/57.0 | 10/9/26/18/88<br>6.6/6.0/17.2/11.9/58.3 | 158 / 194 /830/363 /1653<br>4.9 /6.1/26.0/11.4/51.7 | 118/193/895/376/2057<br>3.2/5.3/24.6/10.3/56.5 |
| Visit to hospital (%) | 7.0 | 31.5*** | 43.9*** | 13.9 | 4.1 |
| Number of symptoms in the first week [median [IQR]] | 5 [3;7] | 7 [5-9]*** | 7 [5;9]*** | 6 [4;8] | 3 [2;4]*** |

\* indicates p <0.1 \*\* <0.05 \*\*\*<0.01 when comparing to short covid. Comparison are performed with respect to the “short duration” within the positive group. Matched Negatives are compared to the overall positive population Mann Whitney U tests are performed for continuous variables and chi square tests are performed when comparing proportions.
Index of Multiple Deprivation (IMD) information is only available for app users from the UK who have entered a complete post code
Acronyms: GB – Great Britain / SE – Sweden / US – United States / IMD – Index of multiple deprivation

**Figure 2:**
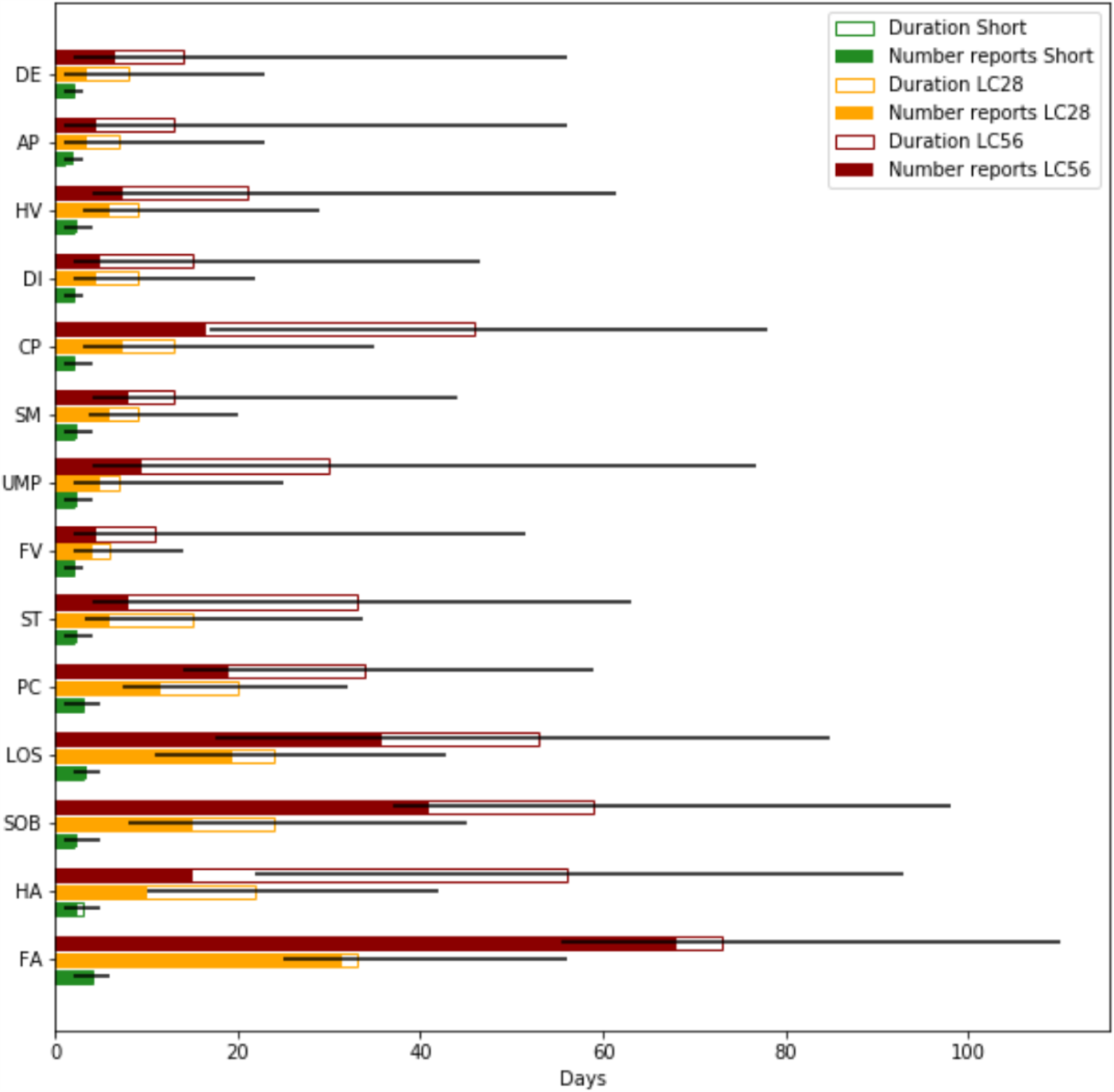
Symptoms by duration. For each symptom (ordered from top to bottom by increasing frequency of occurrence) the median duration of report is presented by the total (hollowed) bar height, with associated interquartile range represented by the black line, for the short, LC28 and LC56 durations. The filled bars represent the number of times a report has been given. For both duration and number of reported days of symptoms, the x axis reflects the number of days. This highlights the differences in the symptoms in terms of their intermittence throughout the course of the disease. (Abbreviations DE – delirium, AP – Abdominal Pain, HV – Hoarse Voice, DI – Diarrhoea, CP – Chest Pain, SM – skipped meals, UMP – Unusual Muscle pains, FV – Fever, ST – Sore Throat, PC – Persistent Cough, LOS – Loss of smell, SOB – Shortness of breath, HA – Headache, FA – Fatigue)

Fatigue (97.7%) and headache (91.2%) were the most reported symptoms in those with LC28, followed by anosmia and lower respiratory symptoms, and headache was more often reported intermittently (Figure 2, supplementary Table s1). Free-text additional symptoms were more commonly reported in LC28 cases (81%) compared to Short-COVID (45%), with cardiac symptoms (palpitations, tachycardia) (LC28,6.1%; short-COVID 0.5% p <0.0005), concentration or memory issues (4.1% vs 0.2%, p<0.0005), tinnitus and earache (3.6% vs 0.2% p<0.0005) and peripheral neuropathy symptoms (pins and needles and numbness) (2% vs 0.5% p=0.004) disproportionately reported in LC28. Most of these symptoms were reported for the first time 3-4 weeks post-symptom onset.

We found two main patterns of symptomatology within LC28: those reporting exclusively fatigue, headache and upper respiratory complaints (shortness of breath, sore throat, persistent cough and loss of smell) and those with additional multi-system complaints, including ongoing fever and gastroenterological symptoms (Supplementary figure 3). In the individuals with long duration (LC28), ongoing fever OR 2.16 [1.50;3.13] and skipped meals OR 2.52 [1.74;3.65] were strong predictors of a hospital visit. Details of the frequency of symptoms persisting beyond 28 and 56 days after disease onset are provided in Supplementary table 3.

**Figure 3:**
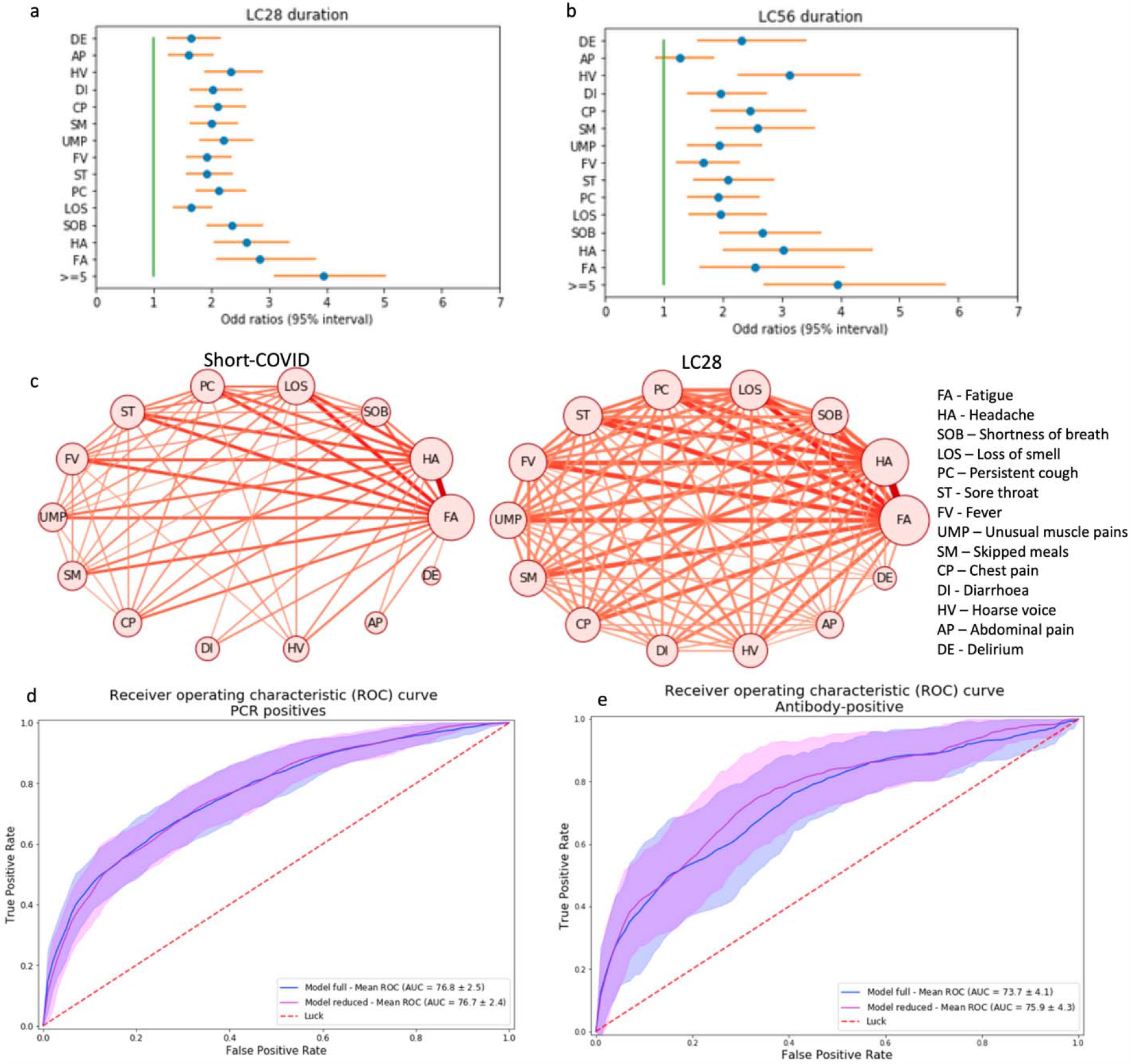
Symptom correlates of long-COVID for LC28 (a) and LC56 (b) with correction for age and gender. c) Co-occurrence network of symptom pairs with the frequency of symptom report as the size of the node and the likelihood of symptom pair co-occurrence by the weight of the edge linking them. Edges representing a co-occurrence of less than 10% were removed. d) – Receiver Operating Characteristic (ROC) curve of the cross-validated full and reduced models on the PCR cohort. e)– ROC curve when training on the whole PCR cohort and testing on the antibody-positive cohort for the full (blue) and reduced (magenta) model. Random predictive probability is indicated in both panels as a dashed red line. (Abbreviations DE – delirium, AP – Abdominal Pain, HV – Hoarse Voice, DI – Diarrhoea, CP – Chest Pain, SM – skipped meals, UMP – Unusual Muscle pains, FV – Fever, ST – Sore Throat, PC – Persistent Cough, LOS – Loss of smell, SOB – Shortness of breath, HA – Headache, FA – Fatigue)

Individuals with LC28 were more likely to report relapses (16.0% vs 8.4%) (p<0.0005). In comparison, in the matched group of SARS-CoV2 negative-tested individuals, relapse was reported in 11.5%, and relapse was longer in LC28 (median = 9 [5-18] vs 6 [4-10] days). We explored how to estimate risk of LC28 among positive individuals from data available early in the disease course. Individuals reporting more than 5 symptoms in the first week (the median number reported) were significantly more likely to go on to experience LC28, (OR=3.95 [3.10;5.04]). This strongest risk factor was predictive in both sexes and all age groups (supplementary Figures 4, a-e).

The five symptoms experienced during the first week most predictive of LC28 in the positive individuals were: fatigue OR=2.83 [2.09;3.83], headache OR=2.62 [2.04;3.37], dyspnoea OR=2.36 [1.91;2.91], hoarse voice OR=2.33 [1.88;2.90] and myalgia OR=2.22 [1.80;2.73] (Figure 3). Similar patterns were observed in both genders. In adults aged over 70, loss of smell (which is less common) was the most predictive of long-COVID OR=7.35 [1.58;34.22] before fever OR=5.51 [1.75;17.36] and hoarse voice OR=4.03[1.21;13.42] (Supplementary figures 4). Co-occurrence plots of symptoms in short-COVID versus LC28 further illustrate the importance of early multi-symptom involvement (Figure 3c).

We created Random Forest Prediction models using a combination of the first week’s symptom reporting, personal characteristics and comorbidities. Using all features, the average ROC AUC was 76.7% (SD=2.5) (Figure 3d) in the classification between short-COVID and LC28. The strongest predictor was age (29.2%) followed by the number of symptoms during the first week (16.3%). Feature importance was relatively similar across age-specific models. However, in the over 70s, early features such as fever, anosmia and comorbidities were important, and may be ‘red flags’ in older adults (Supplementary figure 6).

To create a model usable in healthcare settings, we simplified the prediction model to include only symptom number in the first week with age, and sex in a logistic regression model obtaining ROC AUC of 76.7% (SD 2.5) (Figure 3d), for which the calibration slope had an average of 1.02 (0.15). When optimising the balance between false positives and false negatives, we obtained a specificity of 73.4% (SD 9.7) and a sensitivity of 68.7% (SD 9.9). Specificity, Sensitivity, PPV and NPV values at different thresholds are presented in Supplementary table 6.

Key predictive findings of our analysis were validated in an independent dataset of 2412 individuals who reported testing antibody positive (but no positive PCR result) for SARS-CoV2 from 2 weeks after symptom onset where, again, the number of symptoms in the first week of illness was the strongest predictor, OR=4.60 [95% CI 3.28; 6.46]. The simple prediction model, was similarly predictive of LC28 in the antibody group, with a ROC-AUC of 75.9% (SD=4.3%) (Figure 3-e).

While this study provides important insights into the disease presentation, any generalisation should be considered carefully. Our study was limited by being confined to app users who were disproportionately female and under-represented those >70years which could increase or decrease our estimate of the extent of Long-COVID respectively and caution is needed in interpreting associations found in smaller population subgroups. Swab test results were self-reported and were all assumed to be RT-PCR, as antigen tests were not available at the time. Applying a weighting following the UK population (see Supplementary Methods), the estimated proportion of people experiencing symptomatic COVID-19 going on to suffer Long-COVID were similar: 14.5%, 5.1% and 2.2% for 4, 8- and 12-weeks duration respectively. While estimates could be inflated because early PCR testing was restricted to those more severely unwell, or if regular logging or test results encouraged a systematic bias in symptom reporting, Long-COVID may here be underestimated if individuals with prolonged symptoms were more likely to stop logging symptoms on the app. Our participant selection criteria were chosen to confidently identify cases, and upper and lower bounds for estimates given each exclusion criteria are presented in Supplementary Table 4. Symptom reporting rates through the study period for all users are also presented in Supplementary Table 6. Taken together, these data suggest that our estimates may be conservative. We had insufficient numbers to explore risk factors for disease over 2 months and were unable to analyse the impact of ethnicity due to incomplete data. In addition, the list of symptoms on the app is necessarily non-exhaustive, however, analysis of the free-text responses allowed us to highlight other symptoms present in Long-COVID, such as cardiac and neurological manifestations. With emerging evidence of ongoing myocardial inflammation and change in^8,9^ associated with COVID-19, this calls for specific studies of cardiac and neurological longer-term sequelae of COVID-19.

At the population level, it is critical to quantify the burden of Long-COVID to better assess its impact on the healthcare system and appropriately distribute resources. In our study, prospective logging of a wide range of symptoms allowed us to conclude that the proportion of people with symptomatic COVID-19 who experience prolonged symptoms is considerable, and relatively stable across three countries with different cultures. Whether looking at a four-week or an eight-week threshold for defining long duration, those experiencing Long-COVID were consistently older, more likely to be female and to require hospital assessment than in the group reporting symptoms for a short period of time. Those going on to experience LC28 had multi-system disease from the start, supporting the need for holistic support^10^. While asthma was not reported as a factor of risk for hospitalisation in^11^, its association with Long-COVID (LC28) warrants further investigation.

We found early disease features were predictive of duration. With only three features - number of symptoms in the first week, age and sex, we were able to accurately distinguish individuals with LC28 from those with short duration. Importantly, the model generalised well to the population reporting antibody testing. This important information could feature in highly needed targeted education material for both patients and healthcare providers and we present typical nomograms for use in clinical settings in Supplementary Figure 7. Moreover, the method could help determine at-risk groups and could be used to target early intervention trials and clinical service developments to support rehabilitation in primary and specialist care^14^ to alleviate Long-COVID and facilitate timely recovery.

## Data Availability

Data used in this study is available to bona fide researchers through UK Health Data Research using the following link

https://web.www.healthdatagateway.org/dataset/fddcb382-3051-4394-8436-b92295f14259

## Ethics

In the UK, the App Ethics has been approved by KCL ethics Committee REMAS ID 18210, review reference LRS-19/20-18210 and all subscribers provided consent. In Sweden, ethics approval for the study was provided by the central ethics committee (DNR 2020-01803).

## Data and materials availability

Data used in this study is available to bona fide researchers through UK Health Data Research using the following link https://web.www.healthdatagateway.org/dataset/fddcb382-3051-4394-8436-b92295f14259

## References

1. Wiersinga, W. J., Rhodes, A., Cheng, A. C., Peacock, S. J. & Prescott, H. C. Pathophysiology, Transmission, Diagnosis, and Treatment of Coronavirus Disease 2019 (COVID-19): A Review. JAMA - Journal of the American Medical Association vol. 324 782–793 (2020).

2. Sheehy, L. M. Considerations for postacute rehabilitation for survivors of COVID-19. Journal of Medical Internet Research vol. 22 (2020).

3. Alwan, N. A. A negative COVID-19 test does not mean recovery. Nat. 2020 5847820 (2020).

4. Menni, C. et al. Real-time tracking of self-reported symptoms to predict potential COVID-19. Nat. Med. 26, 1037–1040 (2020).

5. Drew, D. A. et al. Rapid implementation of mobile technology for real-time epidemiology of COVID-19. Science (80-). eabc0473 (2020) doi:10.1126/science.abc0473.

6. Sudre, C. H. et al. Symptom clusters in Covid19: A potential clinical prediction tool from the COVID Symptom study app. medRxiv 2020.06.12.20129056 (2020) doi:10.1101/2020.06.12.20129056.

7. Cirulli, E. T. et al. Long-term COVID-19 symptoms in a large unselected population. medRxiv (2020) doi:10.1101/2020.10.07.20208702.

8. Mitrani, R. D., Dabas, N. & Goldberger, J. J. COVID-19 cardiac injury: Implications for long-term surveillance and outcomes in survivors. Hear. Rhythm (2020) doi:10.1016/j.hrthm.2020.06.026.

9. Padda, I., Khehra, N., Jaferi, U. & Parmar, M. S. The Neurological Complexities and Prognosis of COVID-19. SN Compr. Clin. Med. (2020) doi:10.1007/s42399-020-00527-2.

10. Living with Covid19. https://evidence.nihr.ac.uk/themedreview/living-with-covid19/ (2020) xdoi:10.3310/themedreview_41169.

11. Robinson, L. B. et al. Title: COVID-19 severity in asthma patients: A multi-center matched cohort study. medRxiv 2020.10.02.20205724 (2020) doi:10.1101/2020.10.02.20205724.

12. Mahase, E. Covid-19: Demand for dexamethasone surges as RECOVERY trial publishes preprint. BMJ 369, m2512 (2020).

13. Beigel, J. H. et al. Remdesivir for the Treatment of Covid-19 — Preliminary Report. N. Engl. J. Med. (2020) doi:10.1056/nejmoa2007764.

14. Greenhalgh, T., Knight, M., A’Court, C., Buxton, M. & Husain, L. Management of post-acute covid-19 in primary care. BMJ 370, (2020).

15. Drew, D. A. et al. Rapid implementation of mobile technology for real-time epidemiology of COVID-19. Science (80-.). (2020).

16. Spiel, C. et al. A Euclidean distance-based matching procedure for non randomized comparison studies. Eur. Psychol. 13, 180–187 (2008).

