## Supplemental file for "Attributes and predictors of Long-COVID: analysis of COVID cases and their symptoms collected by the Covid Symptoms Study App"

**Online methods**

**Methods**

**Dataset:**

Data used in this study were acquired through the COVID 19 Symptom Study app, a mobile health application developed by Zoe Global Limited with input from physicians and scientists at King’s College London, Massachusetts General Hospital, Lund and Uppsala Universities^15^. The app, which collects data on personal characteristics and enables prospective logging of symptoms, was launched in the UK, the US and Sweden between 24 March 2020 (UK) and 30 April 2020 (Sweden), and rapidly reached over 4 million users from the community. App users are asked to report their health status daily, and any incident COVID-test (both undertaking of the test and its result). Questions on the app are appended below. The current study focuses on 4182 users who reported testing positive to SARS-CoV2 by PCR swab test with symptom onset between 25 March 2020 and 30 June 2020, for whom the date of symptom onset matched clinically with the date of test and in whom duration of symptoms could be estimated (Supplementary Figure 1 presents a flowchart of study inclusion). We repeated the analyses in an independent subgroup of 2412 app users who reported positive testing for antibodies against SARS-CoV2 at least two weeks after symptom onset, but without swab test results (Supplementary Figure 1).

To understand how the duration and relapse rate compared to a comparable population not suffering from COVID-19, we selected an additional matched sample from all app users meeting study inclusion criteria but who tested negative by PCR swab test, choosing for each COVID+ case the individual from the negative group with the smallest Euclidean distance based on sex, age, and BMI ^16^.

**Definitions**

Onset of disease was defined as the first day of reporting at least one symptom that had lasted more than one day.

Disease end was defined as the last day of symptom reporting before reporting as healthy for the next consecutive seven days; or the last day of reporting with fewer than 5 symptoms before ceasing using the app. For included participants who had ceased using the app whose cumulative number of symptoms were fewer than 5, disease end was considered as the last log.

Relapse was defined as two or more days of symptoms within a 7-day window after one week of healthy logging, if initial symptoms were temporally close to a positive swab test.

Long-COVID was defined as symptoms persisting for more than 4 weeks (28 days, LC28), more than 8 weeks (56 days, LC56). or more than 12 weeks (LC84) between symptom onset and end, while short duration was defined as the interval between symptom onset and end of less than 10 days, without a subsequent relapse (Short-COVID).

**Inclusion/Exclusion criteria**

To be included in the subsequent analysis, users of the COVID Symptoms Study app were selected based on the following criteria:

*Inclusion criteria*: Age >=18 years; BMI greater than 15 and less than 55,

a positive SARS-CoV-2 swab test (PCR) confirming the diagnosis of COVID-19; disease onset between 14 days before and 7 days after the test date, and before the 30 June 2020 (to limit right censoring).

*Exclusion criteria*: individuals who started app reporting when already unwell; users reporting as exclusively healthy throughout the study period; users with gaps of more than seven days after an unhealthy report who did not report any hospital visit (to allow for gaps due to hospitalisation). In addition, individuals reporting for fewer than 28 days but who reported more than five symptoms at their last log were excluded, as symptom duration could not be ascertained.

Inclusion and exclusion criteria were similar for the matched negative-tested sample, which differed only on the result of their RT-PCR test.

In order to assess the impact of the different exclusion criteria on rates of LC28, Supplementary Table 4 presents the lower and upper bounds of these proportions according to lower and upper bounds assumptions on duration. This table also includes the estimation of proportion of LC28 when accounting for a possible rate of false negatives ranging from 2 to 30% based on the distribution estimated from the matched negative sample. Table 5 reflects the demographics of the different excluded groups.

**Statistical testing and modelling**

Data collected prospectively until 02 September were included, to allow sufficient time to ascertain duration. Univariable and multivariable logistic regression was used to assess symptoms associated with short- and long-COVID respectively, adjusting for sex and age, using Statsmodels v0.11.1 Python3.7. Separate models were fitted to subgroups stratified by sex and age (18-49; 50-69; >70 years). For analysis of relapse, existence and duration of relapse were compared between the LC28 group and the whole control sample, using a Mann Whitney U test.

We used a K-mode clustering analysis to investigate whether there was evidence of different sub-types of long-COVID, using the kmode package v0.10.2. Number of ideal symptom clusters was obtained via a silhouette analysis with dice distance metrics. Differences between LC28 and short-COVID were visualised using a co-occurrence network (networkx for visualisation), applying a 10% threshold to remove rare edges to aid visualisation.

Finally, to create a predictive model for Long-COVID LC28, we used sklearn v0.22.2.post1 package, training random forest classifiers with stratified repeated cross-validation (10 times, 5 folds) with hyperparameter grid search including, as features, information available during the first week of illness, reported comorbidities (asthma, lung disease, heart disease, kidney disease and diabetes) and personal characteristics (BMI, age, sex). In addition to a global consideration of the studied sample population, separate models stratified by age were also entrained using a similar cross-validation setting (hyperparameter search and stratified sampling). After running the cross-validation for each model structure (50 times), the feature importance was averaged across the different repeated folds. To create a simplified linear model, we applied a Lasso least angular regression information criterion with Bayesian information criterion was used for feature selection. This resulted in a model that included only age, gender, and the number of symptoms experienced during the first week.

Using only these three features, a logistic regression model was then assessed using the same stratification and cross-validation.

To assess performance on the test dataset (antibody positive), cross-validation was also performed to obtain an indication of the variability in performance using models that were trained on the whole PCR-positive sample.

For the reduced logistic regression model, the score was given by the following formula:

S = 0.259503 * NumberSymptoms	+	0.055457 * Age	-0.633310 * Sex – 3.20 (where sex is encoded as 1 – Female / 2 – Male)

**Matching with negative sample:**

The selected negative cases followed the same inclusion rules and were matched to the positive samples using the minimum Euclidean distance between the vectors of features created by age, BMI, and sex applying an Hungarian matching algorithm. Sex feature was multiplied by 100 to ensure balance between feature strength.

**Rebalancing to UK population demographics**

Lastly, the rebalancing with respect to the UK population was performed by reweighting the age/sex proportions of LC28 in the studied sample by that of the UK population based on census data from 2018. The weighting per age group is described in the table below:

|  | Female | Male |
| --- | --- | --- |
| 18-49 | 0.263 | 0.264 |
| 50-69 | 0.156 | 0.150 |
| >=70 | 0.093 | 0.075 |

**Ascertainment of parameters**

The wording of the questions on the app when registering and when later describing symptom presentation is described below. Specific comments regarding changes and interpretation are in square brackets.

*Ascertainment of demographic characteristics and medical history*

Demographic and medical history assessment

What year were you born?

What sex were you assigned at birth?

- Female
- Male
- Prefer not to say
- Intersex

Your height?

Your weight?

Do you have heart disease?

Do you have diabetes?

    What kind of diabetes do you have? [question added June 19]

- Type 1 diabetes
- Type 2 diabetes
- Gestational diabetes (during pregnancy)
- Unsure
- Prefer not to say
- Other

Do you have lung disease or asthma? - [Changed on 6 June to: the following]

Do you have asthma?

Do you have lung disease?]

Do you have kidney disease?

*Assessment of symptom experience*

Do you have a fever or feel too hot?

Do you have persistent cough (coughing a lot for more than an hour, or 3 or more coughing episodes in 24h?

Are you experiencing unusual fatigue?

- No
- Mild
- Severe (I struggle to get out of bed)

Are you experiencing unusual shortness of breath?

- No
- Yes - Mild symptoms (slight shortness of breath during ordinary activity)
- Yes - Significant symptoms (breathing is comfortable only at rest)
- Yes - Severe symptoms (breathing is difficult even at rest)

Do you have a loss of smell or taste?

Do you have an unusually hoarse voice?

Are you feeling an unusual chest pain or tightness to your chest?

Do you have an unusual abdominal pain?

Are you experiencing diarrhoea?

Do you have a headache?

Have you been skipping meals?

Do you have any of the following symptoms: confusion, disorientation or drowsiness?

Do you have a sore throat?

Do you have unusual strong muscle pains?

*Assessment of test:*

Assessment of test result was modified on the 1^st^ of May. Tests reported until that day are all assumed to be RT-PCR tests. It became possible after that date for the users to specify the type of test.

Initial version (Before 1^st^ May 2020)

Have you had a Covid Test?

Did you test positive for Covid19?

- Yes
- No
- Waiting

From 1st May - When was your test? [If unable to give specific date, question was transformed to]

When was your test roughly?

Second version

How was the test performed?:

- Nose or throat swab [interpreted as PCR]
- Spat in a tube/cup [interpreted as PCR]
- Pricked my finger and gave blood [interpreted as antibody]
- My blood was drawn via a needle [interpreted as antibody]
- Other (specify)

Did you test positive for COVID-19?

- Positive
- Negative
- Failed
- Waiting

**Supplementary Tables**

**Supplementary Table 1:** Comparison for each symptom of the frequency (over the whole disease course), existence in the first week, duration (defined as the difference between first and last day on which a particular symptom was reported) and number of reports of that symptom, for Short-COVID, LC28 and LC56 groups. For comparison purposes similar measures are made for the group of matched negative individuals. Symptoms are ordered by overall frequency in the LC28 group.

|  | **Occurrence overall** | | | | | **Occurrence first week** | | | | | **Duration of symptom (median [IQR])** | | | | | **Number of reports (median [IQR])** | | | | |
| --- | --- | --- | --- | --- | --- | --- | --- | --- | --- | --- | --- | --- | --- | --- | --- | --- | --- | --- | --- | --- |
|  | **PCR positives** | | | | **PCR neg** | **PCR positives** | | | | **PCR neg** | **PCR positives** | | | | **PCR neg** | **PCR positives** | | | | **PCR neg** |
|  | **Short** | **L28** | **L56** | **Overall** |  | **Short** | **L28** | **L56** | **Overall** |  | **Short** | **L28** | **L56** | **Overall** |  | **Short** | **L28** | **L56** | **Overall** |  |
| **FA** | 76.10% | 97.70% | 96.80% | 85.87% | 57.99% | 75.90% | 89.10% | 87.80% | 82.31% | 56.43% | 4 [2 ; 6] | 33 [25 ; 56] | 73 [55.5 ; 110] | 8 [4 ; 15] | 3 [1 ; 6] | 4 [2 ; 6] | 31.5 [19.5 ; 54.5] | 68 [39 ; 118.5] | 8 [4 ; 16] | 3 [1.5 ; 6] |
| **HA** | 65.60% | 91.20% | 93.70% | 77.21% | 57.82% | 65.40% | 80.60% | 81.50% | 73.53% | 55.88% | 3 [1 ; 5] | 22 [10 ; 42] | 56 [22 ; 93] | 6 [2 ; 11] | 2 [1 ; 5] | 2.5 [1.5 ; 4] | 10 [4.5 ; 19.5] | 15 [7 ; 34.5] | 4 [2 ; 7.5] | 2 [1 ; 4] |
| **SOB** | 29.40% | 70.80% | 75.70% | 44.07% | 16.00% | 29.20% | 48.40% | 51.30% | 36.56% | 14.73% | 2 [1 ; 5] | 24 [8 ; 45] | 59 [37 ; 98] | 5 [2 ; 12] | 3 [1 ; 5] | 2.5 [1.5 ; 4.5] | 15 [5.5 ; 34.5] | 41 [15.75 ; 80.25] | 5 [2 ; 10] | 2.5 [1.5 ; 5] |
| **LOS** | 49.20% | 72.00% | 75.10% | 60.33% | 7.34% | 49.00% | 56.80% | 58.20% | 54.35% | 6.70% | 3 [2 ; 5] | 24 [11 ; 42.75] | 53 [17.5 ; 84.75] | 6 [3 ; 11] | 2 [1 ; 5] | 3.5 [2 ; 5] | 19.5 [9 ; 34] | 35.75 [11.63 ; 68.88] | 6 [3 ; 10.5] | 2 [1.5 ; 5] |
| **PC** | 40.50% | 68.60% | 62.40% | 53.73% | 18.87% | 40.40% | 57.00% | 52.90% | 49.19% | 17.55% | 3 [1 ; 5] | 20 [7.5 ; 32] | 34 [14 ; 59] | 6 [2 ; 12] | 3 [1 ; 6] | 3 [1.5 ; 4.5] | 11.5 [4.5 ; 22.5] | 19 [7 ; 40.5] | 5 [2.5 ; 9] | 3 [1.5 ; 5.5] |
| **ST** | 41.20% | 67.00% | 72.50% | 52.56% | 44.86% | 41.20% | 53.60% | 54.00% | 48.68% | 43.35% | 2 [1 ; 4] | 15 [3.25 ; 33.75] | 33 [4 ; 63] | 4 [2 ; 8] | 3 [1 ; 5] | 2.5 [1.5 ; 4] | 6 [3 ; 12] | 8 [3 ; 17] | 3 [2 ; 6] | 2.5 [1.5 ; 4.5] |
| **FV** | 36.10% | 62.90% | 58.70% | 46.53% | 20.99% | 36.10% | 50.50% | 45.50% | 42.75% | 19.73% | 2 [1 ; 3] | 6 [2 ; 14] | 11 [2 ; 51.5] | 3 [1 ; 7] | 2 [1 ; 3] | 2 [1.5 ; 3] | 4 [2 ; 7.75] | 4.5 [2 ; 11.5] | 2.5 [1.5 ; 5] | 2 [1 ; 3] |
| **UMP** | 29.20% | 64.00% | 64.60% | 42.35% | 20.01% | 29.20% | 47.50% | 43.40% | 37.57% | 18.58% | 2 [1 ; 4] | 7 [2 ; 25] | 30 [4 ; 76.75] | 3 [1 ; 7] | 2 [1 ; 4] | 2.5 [1.5 ; 3.5] | 5 [2 ; 11] | 9.5 [3 ; 30.63] | 3 [1.5 ; 6] | 2 [1 ; 3.5] |
| **SM** | 29.90% | 59.50% | 66.70% | 41.77% | 15.90% | 29.90% | 46.60% | 52.40% | 37.33% | 15.21% | 2 [1 ; 4] | 9 [3.75 ; 20] | 13 [4 ; 44] | 4 [2 ; 9] | 2 [1 ; 4] | 2.5 [1.5 ; 4] | 6 [3 ; 14] | 8 [3.5 ; 18.88] | 3.5 [2 ; 6.5] | 2 [1 ; 3.5] |
| **CP** | 28.20% | 60.00% | 63.00% | 40.22% | 17.69% | 28.10% | 42.50% | 45.00% | 34.55% | 16.48% | 2 [1 ; 4] | 13 [3 ; 35] | 46 [17 ; 78] | 4 [1 ; 9] | 2 [1 ; 5] | 2 [1.5 ; 4] | 7.5 [2.5 ; 16.5] | 16.5 [7.25 ; 42] | 3 [1.5 ; 6.5] | 2.5 [1.5 ; 4.5] |
| **DI** | 20.40% | 51.10% | 54.50% | 32.04% | 20.64% | 20.10% | 34.60% | 33.30% | 26.18% | 19.27% | 2 [1 ; 3] | 9 [2 ; 22] | 15 [2 ; 46.5] | 3 [1 ; 7] | 1 [1 ; 4] | 2 [1 ; 3] | 4.5 [2 ; 9] | 5 [2 ; 12.25] | 2.5 [1.5 ; 5] | 2 [1 ; 3] |
| **HV** | 23.00% | 53.00% | 61.40% | 34.67% | 12.96% | 22.90% | 41.40% | 47.60% | 31.04% | 11.93% | 2 [1 ; 4] | 9 [3 ; 29] | 21 [4 ; 61.5] | 4 [1 ; 8] | 2 [1 ; 4] | 2.5 [1.5 ; 4] | 6 [3 ; 14.5] | 7.5 [3.5 ; 20.625] | 3.5 [2 ; 6.5] | 2.5 [1.5 ; 4] |
| **AP** | 17.20% | 44.10% | 49.20% | 25.25% | 20.85% | 17.00% | 26.00% | 22.20% | 20.66% | 19.54% | 1 [1 ; 3] | 7 [1 ; 23] | 13 [1 ; 56] | 2 [1 ; 6] | 2 [1 ; 4] | 2 [1 ; 3] | 3.5 [1.5 ; 8.5] | 4.5 [2 ; 9.5] | 2.5 [1.5 ; 4.5] | 2 [1 ; 3.5] |
| **DE** | 11.80% | 30.30% | 38.60% | 17.50% | 9.64% | 11.70% | 19.00% | 24.30% | 14.80% | 8.92% | 2 [1 ; 3] | 8 [1 ; 23] | 14 [2 ; 56] | 3 [1 ; 7] | 1 [1 ; 3] | 2 [1 ; 3.5] | 3.5 [1.5 ; 9] | 6.5 [2 ; 16.5] | 2.5 [1.5 ; 5] | 1.5 [1 ; 3] |

**Supplementary Table 2:** Odds ratios [95% CI] for LC28 and LC56 per decade, compared to 18-30years category, and separating males and females.

|  | Females | | Males | |
| --- | --- | --- | --- | --- |
|  | LC28 | LC56 | LC28 | LC56 |
| 30-40 years | 2.11 [1.38 ; 3.23] | 2.19 [1.02 ; 4.73] | 2.62 [1.09 ; 6.27] | 4.12 [0.49 ; 34.67] |
| 40-50 years | 4.35 [2.9 ; 6.53] | 4.14 [2.02 ; 8.52] | 2.24 [0.9 ; 5.62] | 3.52 [0.39 ; 31.91] |
| 50-60 years | 8.03 [5.37 ; 12] | 8.61 [4.33 ; 17.1] | 6.65 [2.8 ; 15.8] | 11.49 [1.44 ; 91.29] |
| 60-70years | 6.53 [4.02 ; 10.6] | 7.2 [3.31 ; 15.69] | 7.5 [2.98 ; 18.9] | 14 [1.68 ; 116.51] |
| >=70 years | 5.46 [2.51 ; 11.85] | 7.71 [2.6 ; 22.83] | 8.27 [2.8 ; 24.39] | 18.56 [1.99 ; 173.3] |

**Supplementary Table 3:** Indication of the percentage of individual ongoing symptoms beyond 28 days and beyond 56 days in the LC28 and the LC56 groups. Abbreviations: FA – Fatigue; HA – Headache; SOB – Shortness of breath; LOS – Loss of smell; PC – Persistent cough; ST – Sore throat; FV – Fever; UMP – Unusual muscle pains; SM – Skipped meals; CP – Chest pain; DI – Diarrhoea; HV – Hoarse voice; AP – Abdominal pain; DE – Delirium.

|  | Reported beyond 28 days in LC28 | Reported beyond 56 days in LC56 |
| --- | --- | --- |
| **FA** | 0.68 | 0.73 |
| **HA** | 0.40 | 0.53 |
| **SOB** | 0.37 | 0.48 |
| **LOS** | 0.39 | 0.39 |
| **PC** | 0.27 | 0.22 |
| **ST** | 0.27 | 0.27 |
| **FV** | 0.12 | 0.16 |
| **UMP** | 0.20 | 0.30 |
| **SM** | 0.13 | 0.19 |
| **CP** | 0.23 | 0.31 |
| **DI** | 0.15 | 0.20 |
| **HV** | 0.17 | 0.22 |
| **AP** | 0.15 | 0.23 |
| **DE** | 0.11 | 0.15 |

**Supplementary Table 4**: Demographic characteristics for individuals meeting major exclusion criterion according to the selection flowchart Figure 1. The first three columns relate to individuals who were asymptomatic (or only one day of symptoms) around the time of the test. Statistical comparisons are made to the final kept sample using chi-square test for frequency measurements and MannWhitney-U test for non normally distributed continuous variables. Continuous variables are presented with median [1^st^ quartile; 3^rd^ quartile]

|  | **Asymptomatic** | | | **Symptomatic** | | | | | |
| --- | --- | --- | --- | --- | --- | --- | --- | --- | --- |
| **Exclusion reason** | No reported symptom | No symptom reported up to 7 days post test | 1 day of symptom only | Starting unhealthy | Symptoms too early^2^ | Not regular logging^3^ | Disease start post censoring date | Dropping^4^ | **Kept** |
| **Number** | 5296 | 4376 | 1539 | 6695 | 1114 | 1652 | 1068 | 470 | **4182** |
| **Asthma (%)** | 8.3*** | 15.2 *** | 8.5 | 9.1 | 12.5 * | 12.0 * | 12.6 * | 3.0 *** | **10.0** |
| **Lung disease (%)** | 9.6*** | 10.3 | 9.5 ** | 13 | 17.1 ** | 15.3 | 11.6 | 13 | **13.6** |
| **Diabetes (%)** | 3.5 | 3.9 | 2.7 | 3.5 | 3.4 | 2.9 | 2.8 | 5.1 * | **2.9** |
| **Male (%)** | 42.1*** | 28.0 | 36.0*** | 29.1 | 28 | 24.7 ** | 33.5 ** | 21.7 ** | **28.5** |
| **Heart (%)** | 3.3*** | 4.6 | 2.4 | 2.5 | 2.7 | 1.8 | 2.6 | 2.1 | **2.0** |
| **BMI (kg/m2)** | 25.9 [23.0;30.0] | 26.8 [23.4;31.5] | 25.8 [22.7;30.4]*** | 26.4 [23.2;31.1] ** | 26.3[22.9;31.1] | 26.5 [23.2;31.0]* | 25.8[22.9;30.0]* | 27.4[23.8;32.6] *** | **25.9 [23.1;30.2]** |
| **Age (years)** | 45 [32;56]*** | 48 [37;57] *** | 36 [27;48] | 41 [32;52] ** | 40 [31;52] * | 41 [31;51] ** | 46 [33;57] *** | 37 [29;49]*** | **42 [32;53]** |
| **NumbSymp^1^** | NA | NA | NA | 7 [4-9] *** | 4 [2-6]*** | 7 [5;9] *** | 5 [3;7] *** | 9 [7;10] *** | **6 [4;8]** |

**^1^** For individuals starting unhealthy at first log, the number of symptoms during first week calculated only for those with entries less than 7 days after the test (only 4876 individuals)

^2^ Test occurring more than 2 weeks after onset of symptoms

^3^(no hospital visit and more than 7 days with missing input after symptomatic logging)

^3^ stopped logging with more than 5 symptoms and duration less than 28 days

Indication of significance when comparing to the cohort kept for the analysis: * p<0.05, ** p<0.01, *** p<0.001

**Supplementary Table 5**: Lower bound and upper bound of estimation of the percentage of individuals with LC28 according to the different criteria of exclusion in the study. Estimation of lower bounds and upper bounds are estimated depending on the associated estimation of duration:

- When starting not healthy, the duration is calculated as upper bound by adding the recorded time of symptom before entering to the app to the duration calculated from logging alone. Lower bound is calculated from symptom logging alone
- When the test date is too far from the onset of symptoms (more than 14 days post onset), the lower bound of duration is calculated considering the test as onset while the upper bound of duration is calculated from symptom logging alone
- When the logging is not regular, with more than 7 days of absent logging after a symptomatic log, lower bound duration is calculated taking the last logged symptom before the gap as the last day of illness while the upper bound is defined based on the next occurrence of a healthy report

In this table, the rows indicate additional criteria of selection. While the first column corresponds to the executed selection, the following columns allow for comparison when one of the criteria is missing.

Adjustment for false negatives in the matched sample is further stated in the first column.

| **Proportion with LC 28 (%)** | **Symptomatic PCR+ with no exclusions** | **Analysis of ONLY individuals starting unhealthy** | **Analysis of ONLY individuals with symptoms onset >14 days before PCR test** | **Analysis of ONLY individuals with no further log for more than seven days after a symptom report (and no report of hospital visit)** |
| --- | --- | --- | --- | --- |
| **Symptomatic PCR+ with no exclusions** | 18.7 - 38.5 | 21.7 - 46.8 | / | / |
| **Start healthy** | 16.4 - 32.1 |  | 26.5 - 54.7 | / |
| **Symptom onset within reasonable period of test** | 15.7 - 28.8 | 20.3 - 42.7 | / | 34.2 - 56.8 |
| **No gap of more than 7 days between symptom reporting** | 10.9 - 20.8 | 12.7 - 33.2 | 24.2 - 32.7 | / |
| **Start before 30th June** | 11.9 - 20.8 | 12.8 - 32.4 | 24.7 - 33.4 | 35.3 - 58.5 |
| **Possible to estimate end (resolution or final log with less than 5 symptoms)** | 13.2 | 15.0 - 20.7 | 25.8 - 31.9 | 36.8 - 56.9 |
| **Demographic variables (age/sex/BMI) available** | **13.3** | 15.3 - 21.7 | 26.1 - 31.5 | 36.9 - 57.2 |
| **Correction when including FN matched sample (2 - 29%)** | 10.2 - 13.0 | / | / | / |

**Supplementary Table 6:** Baseline symptomatology of the app participants over the course of the analysis for PCR positive, PCR negatives and never tested individuals. Individuals reporting antibody test result are not included in this analysis.

|  | **Never PCR tested prior to census date and reporting on census date** | | **PCR tested negative prior to census date and reporting on census date** | | **PCR tested positive prior to census date and reporting on census date** | |
| --- | --- | --- | --- | --- | --- | --- |
|  | **Number** | **Average Number of symptoms** | **Number** | **Average number of symptoms** | **Number** | **Average Number of symptoms** |
| **1st April 2020** | 925346 | 0.465 | 2973 | 1.063 | 1060 | 3.050 |
| **1st May 2020** | 1165634 | 0.113 | 21012 | 0.947 | 3477 | 1.956 |
| **1st June 2020** | 862377 | 0.058 | 64401 | 0.262 | 3369 | 0.987 |
| **1st July 2020** | 826786 | 0.057 | 112052 | 0.271 | 3526 | 0.885 |
| **1st August 2020** | 730438 | 0.038 | 155512 | 0.162 | 3521 | 0.843 |
| **1st September 2020** | 566270 | 0.051 | 165248 | 0.181 | 2978 | 0.867 |

**Supplementary table 7:**

Comparison of PPV, NPV, Sensitivity, Specificity for different logistic regression models using different combinations of the chosen features: a) combination of age, gender and number of symptoms reported over the first week. B) combination of age, gender and one of the 5 symptoms considered as most predictive of LC28

| **Model variables** | **Threshold** | **0.1** | **0.2** | **0.3** | **0.4** | **0.5** | **0.6** | **0.7** | **0.8** | **0.9** |
| --- | --- | --- | --- | --- | --- | --- | --- | --- | --- | --- |
| Age | Specificity | 0.000 | 0.005 | 0.236 | 0.483 | **0.668** | 0.812 | 0.930 | 0.987 | 0.998 |
|  | Sensitivity | 1.000 | 0.999 | 0.932 | 0.813 | **0.639** | 0.383 | 0.135 | 0.023 | 0.005 |
|  | PPV | 0.260 | 0.260 | 0.300 | 0.356 | **0.404** | 0.418 | 0.403 | 0.388 | 0.419 |
|  | NPV | NaN | 0.970 | 0.910 | 0.881 | **0.841** | 0.790 | 0.754 | 0.742 | 0.741 |
| Gender | Specificity | 0.000 | 0.000 | 0.000 | 0.327 | **0.327** | 1.000 | 1.000 | 1.000 | 1.000 |
|  | Sensitivity | 1.000 | 1.000 | 1.000 | 0.798 | **0.798** | 0.000 | 0.000 | 0.000 | 0.000 |
|  | PPV | 0.260 | 0.260 | 0.260 | 0.294 | **0.294** | NaN | NaN | NaN | NaN |
|  | NPV | NaN | NaN | NaN | 0.821 | **0.821** | 0.740 | 0.740 | 0.740 | 0.740 |
| NumbSymp | Specificity | 0.000 | 0.007 | 0.216 | 0.471 | **0.728** | 0.818 | 0.939 | 0.989 | 1.000 |
|  | Sensitivity | 1.000 | 0.994 | 0.916 | 0.790 | **0.556** | 0.436 | 0.206 | 0.042 | 0.000 |
|  | PPV | 0.260 | 0.260 | 0.291 | 0.344 | **0.418** | 0.457 | 0.543 | 0.572 | NaN |
|  | NPV | NaN | 0.706 | 0.881 | 0.865 | **0.824** | 0.805 | 0.771 | 0.746 | 0.740 |
| Age + Gender | Specificity | 0.000 | 0.071 | 0.252 | 0.518 | **0.683** | 0.813 | 0.920 | 0.983 | 0.998 |
|  | Sensitivity | 1.000 | 0.985 | 0.917 | 0.807 | **0.651** | 0.448 | 0.182 | 0.031 | 0.005 |
|  | PPV | 0.260 | 0.271 | 0.301 | 0.371 | **0.420** | 0.459 | 0.448 | 0.391 | 0.452 |
|  | NPV | 1.000 | 0.936 | 0.897 | 0.885 | **0.848** | 0.808 | 0.762 | 0.743 | 0.741 |
| Age + NumbSymp | Specificity | 0.041 | 0.207 | 0.386 | 0.568 | **0.707** | 0.834 | 0.919 | 0.976 | 0.996 |
|  | Sensitivity | 0.998 | 0.963 | 0.896 | 0.795 | **0.671** | 0.547 | 0.365 | 0.187 | 0.043 |
|  | PPV | 0.268 | 0.299 | 0.339 | 0.393 | **0.445** | 0.537 | 0.615 | 0.735 | 0.792 |
|  | NPV | 0.989 | 0.941 | 0.914 | 0.888 | **0.860** | 0.840 | 0.805 | 0.774 | 0.748 |
| Gender + NumbSymp | Specificity | 0.000 | 0.048 | 0.255 | 0.455 | **0.676** | 0.853 | 0.940 | 0.990 | 1.000 |
|  | Sensitivity | 1.000 | 0.988 | 0.906 | 0.779 | **0.616** | 0.413 | 0.215 | 0.047 | 0.000 |
|  | PPV | 0.260 | 0.267 | 0.300 | 0.334 | **0.400** | 0.498 | 0.567 | 0.648 | 0.000 |
|  | NPV | NaN | 0.937 | 0.888 | 0.856 | **0.834** | 0.806 | 0.774 | 0.748 | 0.740 |
| **Age + Gender + NumbSymp** | Specificity | 0.054 | 0.220 | 0.415 | 0.568 | **0.710** | 0.835 | 0.921 | 0.971 | 0.997 |
|  | Sensitivity | 0.998 | 0.956 | 0.891 | 0.788 | **0.671** | 0.545 | 0.378 | 0.209 | 0.051 |
|  | PPV | 0.270 | 0.301 | 0.349 | 0.390 | **0.448** | 0.539 | 0.631 | 0.720 | 0.870 |
|  | NPV | 0.988 | 0.935 | 0.916 | 0.885 | **0.861** | 0.840 | 0.809 | 0.778 | 0.750 |

| **Model variables** | **Threshold** | **0.1** | **0.2** | **0.3** | **0.4** | **0.5** | **0.6** | **0.7** | **0.8** | **0.9** |
| --- | --- | --- | --- | --- | --- | --- | --- | --- | --- | --- |
| Fatigue + Age + Gender | Specificity | 0.020 | 0.131 | 0.282 | 0.526 | **0.691** | 0.818 | 0.914 | 0.979 | 0.998 |
|  | Sensitivity | 0.999 | 0.968 | 0.910 | 0.812 | **0.675** | 0.500 | 0.248 | 0.055 | 0.007 |
|  | PPV | 0.263 | 0.281 | 0.308 | 0.376 | **0.434** | 0.492 | 0.508 | 0.485 | 0.626 |
|  | NPV | 0.987 | 0.924 | 0.901 | 0.889 | **0.858** | 0.823 | 0.776 | 0.747 | 0.741 |
| Headache + Age + Gender | Specificity | 0.016 | 0.140 | 0.296 | 0.517 | **0.692** | 0.820 | 0.919 | 0.979 | 0.999 |
|  | Sensitivity | 0.998 | 0.968 | 0.912 | 0.802 | **0.668** | 0.494 | 0.273 | 0.061 | 0.000 |
|  | PPV | 0.262 | 0.283 | 0.313 | 0.368 | **0.432** | 0.491 | 0.545 | 0.517 | 0.071 |
|  | NPV | 0.966 | 0.928 | 0.906 | 0.882 | **0.856** | 0.822 | 0.783 | 0.748 | 0.740 |
| Shortness of breath + A + G | Specificity | 0.008 | 0.128 | 0.355 | 0.507 | **0.683** | 0.818 | 0.916 | 0.976 | 0.998 |
|  | Sensitivity | 0.998 | 0.980 | 0.910 | 0.815 | **0.669** | 0.445 | 0.257 | 0.091 | 0.013 |
|  | PPV | 0.261 | 0.283 | 0.331 | 0.367 | **0.426** | 0.464 | 0.519 | 0.580 | 0.713 |
|  | NPV | 0.936 | 0.951 | 0.918 | 0.887 | **0.855** | 0.808 | 0.778 | 0.754 | 0.743 |
| Unusual muscle pains + Age+ Gender | Specificity | 0.006 | 0.104 | 0.334 | 0.509 | **0.690** | 0.819 | 0.918 | 0.981 | 0.999 |
|  | Sensitivity | 0.999 | 0.977 | 0.908 | 0.800 | **0.663** | 0.462 | 0.250 | 0.077 | 0.007 |
|  | PPV | 0.260 | 0.277 | 0.323 | 0.364 | **0.429** | 0.473 | 0.520 | 0.586 | 0.672 |
|  | NPV | 0.921 | 0.933 | 0.912 | 0.879 | **0.854** | 0.813 | 0.777 | 0.752 | 0.742 |
| Hoarse voice + Age + Gender | Specificity | 0.002 | 0.098 | 0.325 | 0.507 | **0.688** | 0.829 | 0.926 | 0.974 | 0.999 |
|  | Sensitivity | 1.000 | 0.979 | 0.904 | 0.799 | **0.654** | 0.448 | 0.257 | 0.095 | 0.007 |
|  | PPV | 0.260 | 0.276 | 0.320 | 0.363 | **0.424** | 0.481 | 0.554 | 0.570 | 0.583 |
|  | NPV | 0.948 | 0.933 | 0.907 | 0.879 | **0.850** | 0.811 | 0.780 | 0.754 | 0.741 |
| **Age + Gender + NumbSymp** | Specificity | 0.054 | 0.220 | 0.415 | 0.568 | **0.710** | 0.835 | 0.921 | 0.971 | 0.997 |
|  | Sensitivity | 0.998 | 0.956 | 0.891 | 0.788 | **0.671** | 0.545 | 0.378 | 0.209 | 0.051 |
|  | PPV | 0.270 | 0.301 | 0.349 | 0.390 | **0.448** | 0.539 | 0.631 | 0.720 | 0.870 |
|  | NPV | 0.988 | 0.935 | 0.916 | 0.885 | **0.861** | 0.840 | 0.809 | 0.778 | 0.750 |

**Supplementary Figures**

**Supplementary Figure 1:** Study inclusion criteria.

Individuals reporting symptoms for at most 1 day were considered for the purpose of this analysis to be asymptomatic. We further excluded users who joined the app already unhealthy, for which the onset of disease was not calculable. Of the remainder, we excluded those who only reported intermittent unhealthy report and restricted to individuals reporting prospective symptoms at least once a week over the course of the disease. The left side of the diagram represents the inclusion flowchart for individuals reporting a positive swab test while the right side reflects the inclusion flowchart for individuals with antibody positive test only.

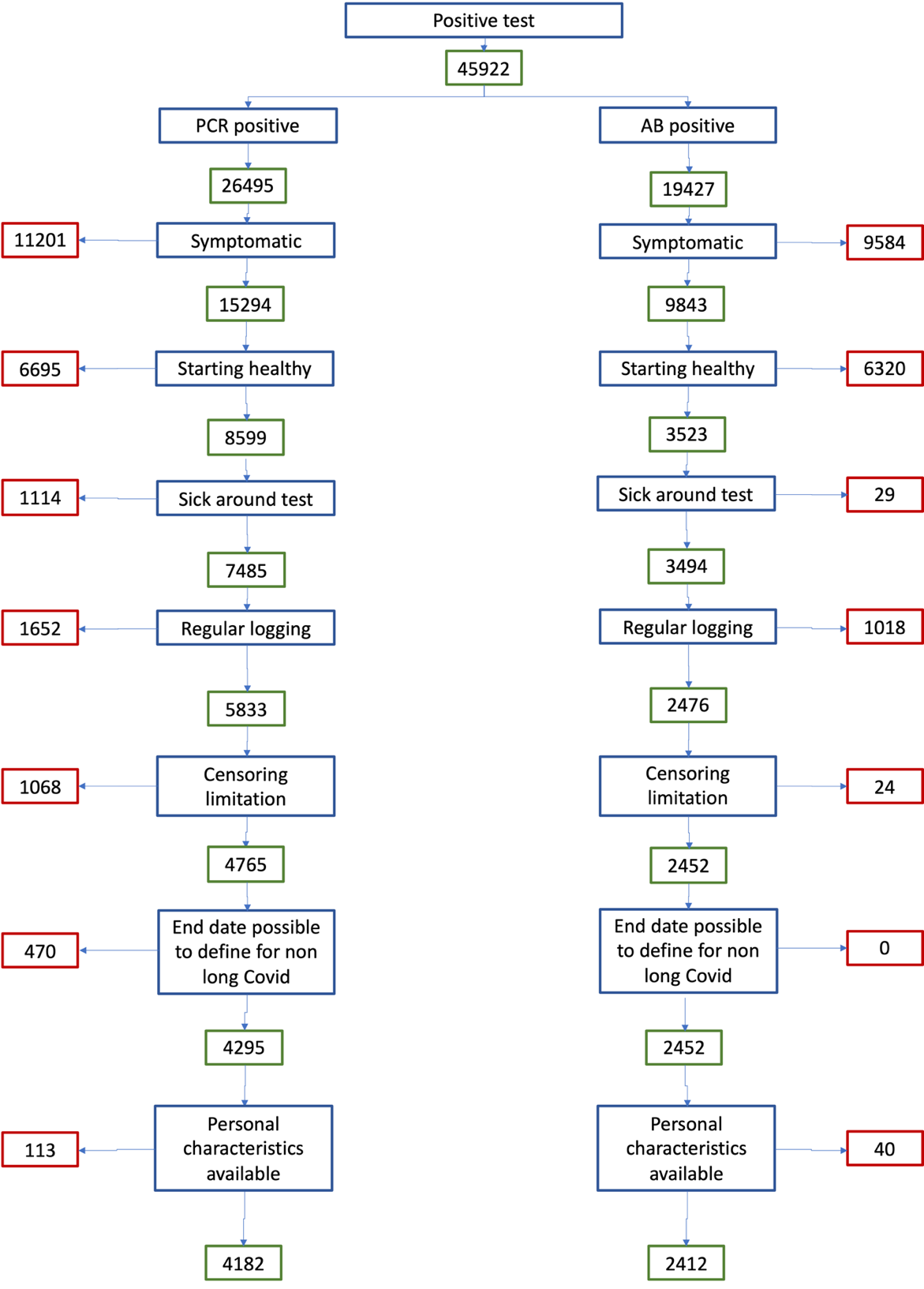

**Supplementary Figure 2:** Ratio of LC28 and LC56 vs short-COVID by IMD quintile

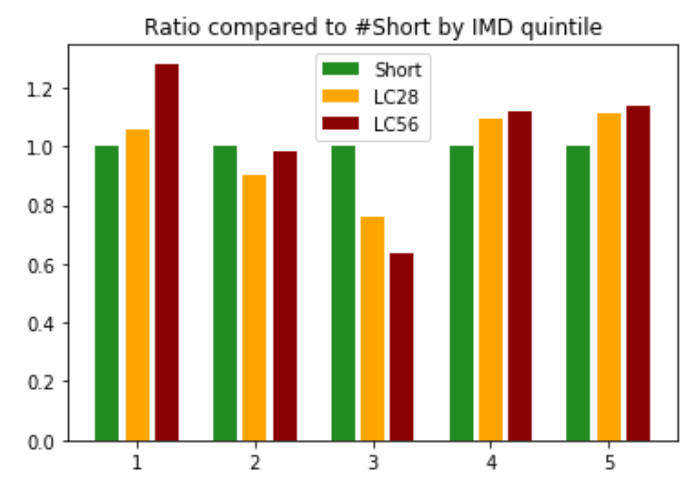

**Supplementary Figure 3** – Clustering of symptoms in the LC28 group, indicating a common strong higher airways component with fatigue, headache, and loss of smell for both groups; and a more multi system presentation for the second group. Colouring presents the frequency of reporting of a given symptom. Abbreviations: DE – delirium, AP – Abdominal Pain, HV – Hoarse Voice, DI – Diarrhoea, CP – Chest Pain, SM – skipped meals, UMP – Unusual Muscle pains, FV – Fever, ST – Sore Throat, PC – Persistent Cough, LOS – Loss of smell, SOB – Shortness of breath, HA – Headache, FA – Fatigue

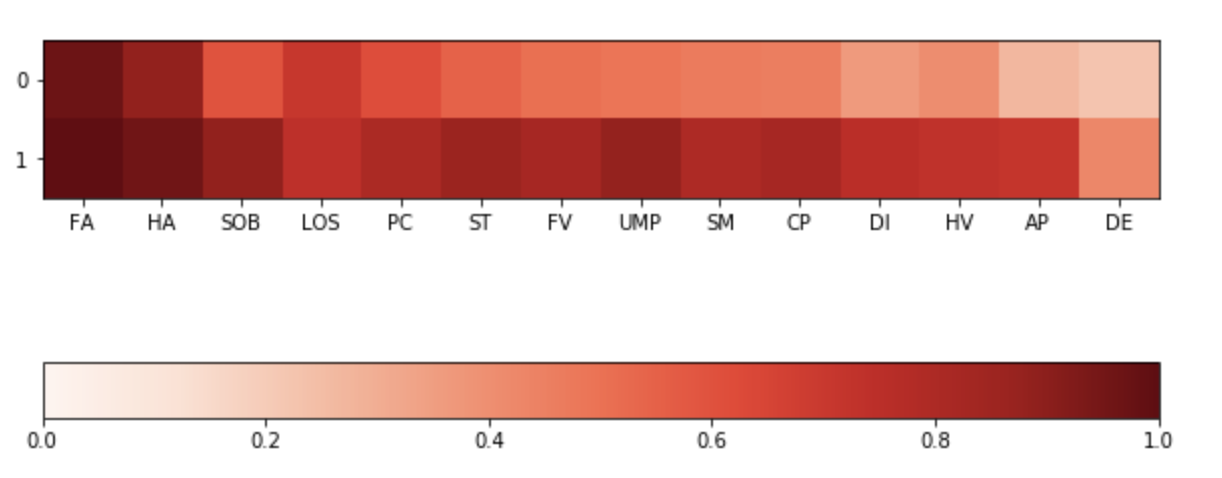

**Supplementary Figure 4.** Odds ratios of LC28 when presenting a given symptom during the first week, correcting for age and gender (if necessary) in different subgroups female(a), male (b), 18-49 (c), 50-69 (d), >=70 (e). Abbreviations: DE – delirium, AP – Abdominal Pain, HV – Hoarse Voice, DI – Diarrhoea, CP – Chest Pain, SM – skipped meals, UMP – Unusual Muscle pains, FV – Fever, ST – Sore Throat, PC – Persistent Cough, LOS – Loss of smell, SOB – Shortness of breath, HA – Headache, FA – Fatigue

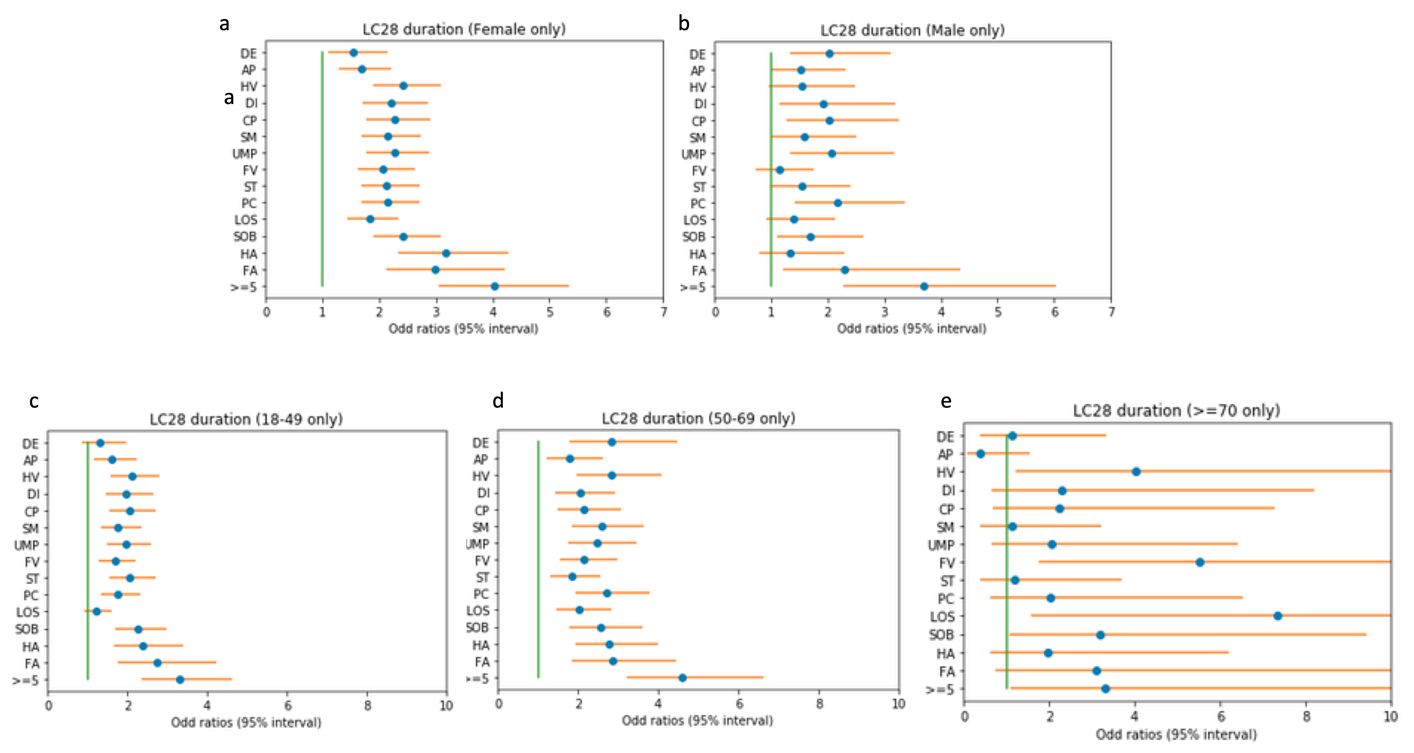

**Supplementary Figure 5** – Odds ratios for the risk of developing Long Covid 28 for each comorbidity or risk factor, correcting for age and gender in each age group,

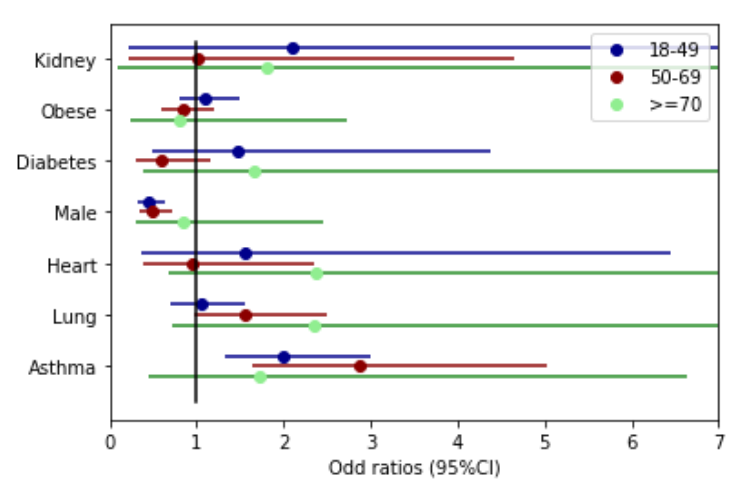

**Supplementary Figure 6:** Comparison of mean feature importance (proportion ranging from 0 to 1) for the cross-validated random forest models across the different age groups when considering personal characteristics and presented symptoms during the first week of the disease. Abbreviations - (Abbreviations DE – delirium, AP – Abdominal Pain, HV – Hoarse Voice, DI – Diarrhoea, CP – Chest Pain, SM – skipped meals, UMP – Unusual Muscle pains, FV – Fever, ST – Sore Throat, PC – Persistent Cough, LOS – Loss of smell, SOB – Shortness of breath, HA – Headache, FA – Fatigue)

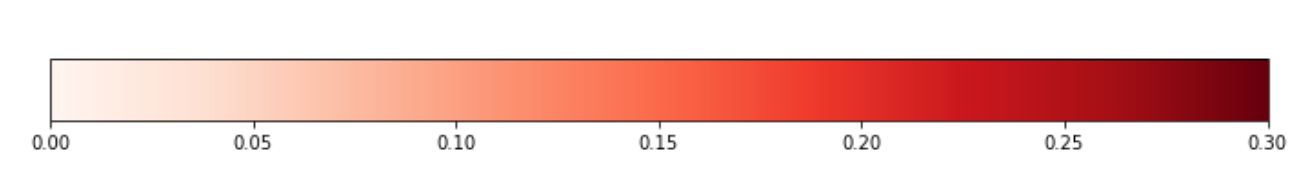

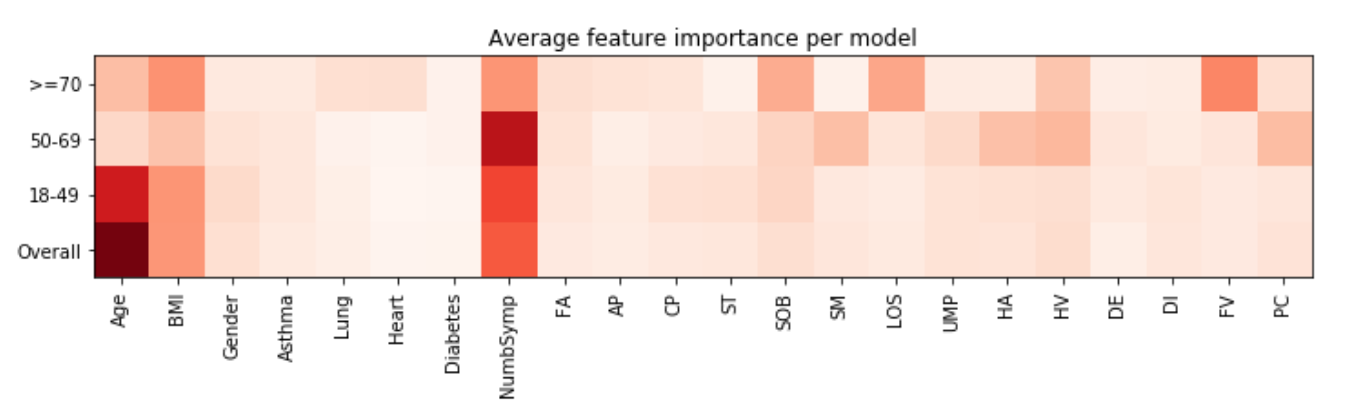

Supplementary Figure 7

Example of nomograms that could be used to assess risk of developing LC28 based on 7 days of symptoms

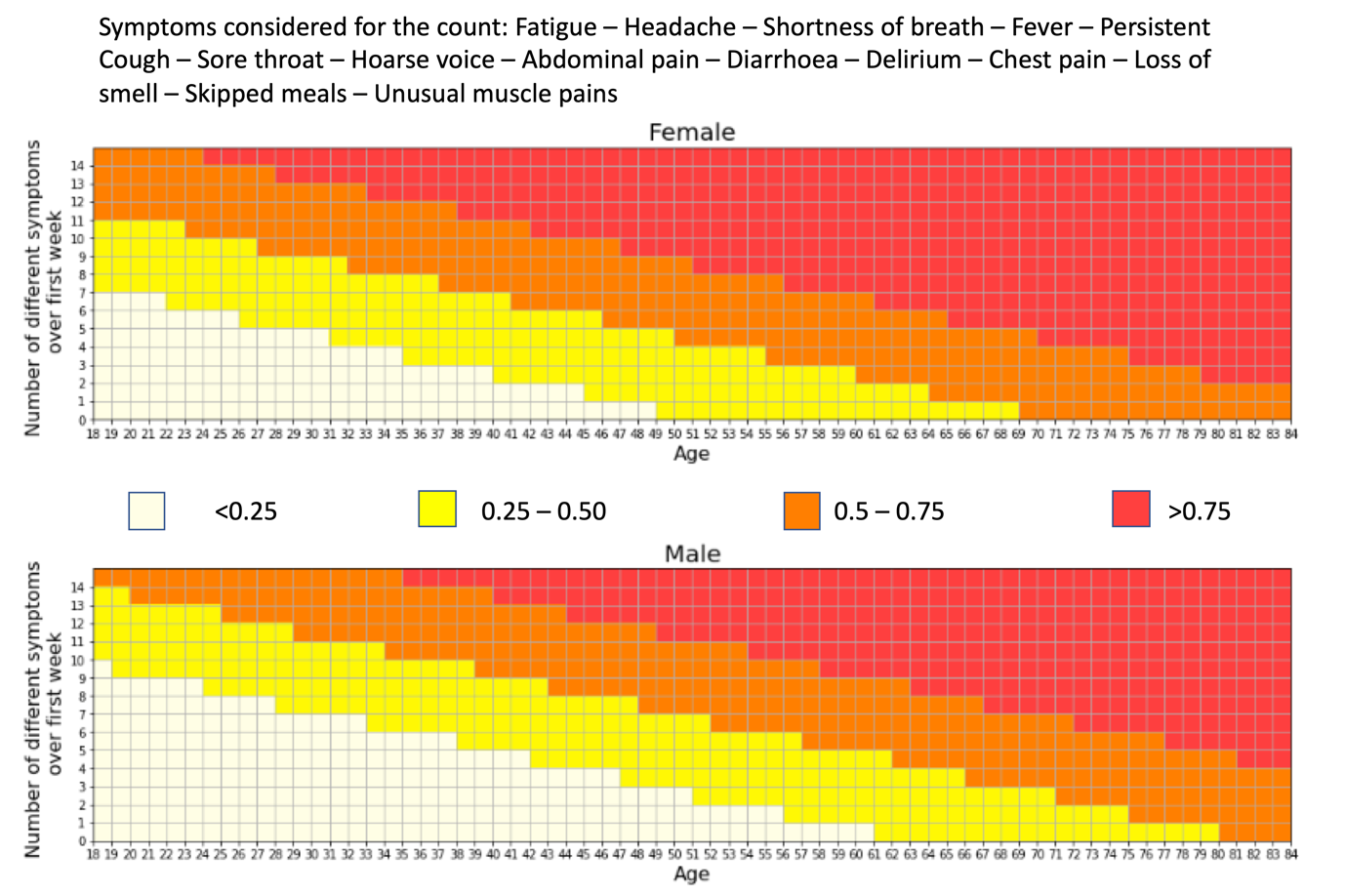
